# Performance evaluation of the Roche Elecsys Anti-SARS-CoV-2 S immunoassay

**DOI:** 10.1101/2021.03.02.21252203

**Authors:** Elena Riester, Peter Findeisen, J. Kolja Hegel, Michael Kabesch, Andreas Ambrosch, Christopher M Rank, Florina Langen, Tina Laengin, Christoph Niederhauser

**Author notes:** Corresponding author Christoph Niederhauser, Interregionale Blood Transfusion Swiss Red Cross, Murtenstrasse 133, 3008 Bern, Switzerland.

## Abstract

**Background:** The Elecsys^®^ Anti-SARS-CoV-2 S immunoassay (Roche Diagnostics International Ltd, Rotkreuz, Switzerland) has been developed for the *in vitro* quantitative detection of antibodies to the severe acute respiratory syndrome coronavirus 2 (SARS-CoV-2) spike (S) protein. We evaluated the performance of this assay using samples from seven sites in Germany, Austria, and Switzerland.

**Methods:** Anonymized frozen, residual serum, or plasma samples from blood donation centers or routine diagnostic testing were used for this study. For specificity and sensitivity analyses, presumed negative samples collected before October 2019 and SARS-CoV-2 PCR-confirmed single or sequential samples were tested, respectively. The performance of the Elecsys Anti-SARS-CoV-2 S immunoassay was also compared with other commercial immunoassays.

**Results:** The overall specificity (n=7880 pre-pandemic samples) and sensitivity (n=240 PCR-positive samples [≥14 days post-PCR]) for the Elecsys Anti-SARS-CoV-2 S immunoassay were 99.95% (95% confidence interval [CI]: 99.87–99.99) and 97.92% (95% CI: 95.21– 99.32), respectively. Compared with seven other immunoassays, the Elecsys Anti-SARS-CoV-2 S assay had comparable or greater specificity and sensitivity. The Elecsys Anti-SARS-CoV-2 S immunoassay had significantly higher specificity compared with the LIAISON^®^ SARS-CoV-2 S1/S2 IgG, ADVIA Centaur^®^ SARS-CoV-2 Total, ARCHITECT SARS-CoV-2 IgG, iFlash-SARS-CoV-2 IgM, and EUROIMMUN Anti-SARS-CoV-2 IgG and IgA assays, and significantly higher sensitivity (≥14 days post-PCR) compared with the ARCHITECT SARS-CoV-2 IgG, iFlash-SARS-CoV-2 IgG and IgM, and EUROIMMUN Anti-SARS-CoV-2 IgG assays.

**Conclusion:** The Elecsys Anti-SARS-CoV-2 S assay demonstrated a robust and favorable performance across samples from multiple European sites, with a very high specificity and sensitivity for the detection of anti-S antibodies.

## Introduction

In December 2019 a novel coronavirus emerged (1), named severe acute respiratory syndrome coronavirus 2 (SARS-CoV-2), which is the causative agent of the disease, COVID-19 (2, 3). SARS-CoV-2 is an enveloped, single-stranded RNA virus of the family Coronaviridae; its genome encodes 16 nonstructural proteins and four structural proteins: spike (S), envelope (E), membrane (M), and nucleocapsid (N) (4). The most prominent protein component on the viral surface is the S glycoprotein – a large transmembrane protein that assembles into trimers to form the distinctive surface spikes of coronaviruses (5, 6). Each S monomer consists of two subunits, S1 and S2, which mediate receptor binding (via the receptor-binding domain [RBD] located in S1) and membrane fusion, respectively, leading to entry into host cells (6-8).

Following infection with SARS-CoV-2, the host mounts an immune response against the virus, including production of specific antibodies against viral antigens (9). Understanding the dynamics of the antibody response to the virus is critical in establishing a relevant time window to use for serology testing (9). Studies into the kinetics of antibodies to SARS-CoV-2 are rapidly emerging and, based on current evidence, both immunoglobulin M (IgM) and G (IgG) antibodies have been detected as early as day 0 to 5 after symptom onset (10, 11). The chronological order of appearance and levels of IgM and IgG appears to be highly variable and often simultaneous (12-14). Several studies have observed median seroconversion at day 10–13 after symptom onset for IgM and day 12–15 for IgG, with maximum seroconversion for IgM, IgG, and total antibodies occurring at week 2–3, week 2– 4, and around week 2, respectively (13-16).

Emergence of the COVID-19 pandemic has resulted in an urgent and unmet need to develop reliable serological tests to determine past exposure to the virus and the seroprevalence in a given population (17). This information is crucial to support diagnosis, contact tracing, epidemiology studies, and vaccine development to enable characterization of pre-vaccination immune status and vaccine-induced immune response (9, 18-20). There are currently 242 candidate SARS-CoV-2 vaccines in development (21) and, of these, 10 are currently in early, limited, or fully approved use (status February 09, 2021) (22). The majority of the vaccines in use are based on the S protein, with the goal of eliciting protective neutralizing antibodies; the rest are based on whole inactivated SARS-CoV-2 (23, 24). Serology assays are also needed for the identification of neutralizing antibodies from convalescent plasma donors (25).

The Elecsys^®^ Anti-SARS-CoV-2 S (Roche Diagnostics International Ltd, Rotkreuz, Switzerland) is an electrochemiluminescence immunoassay (ECLIA), which has been developed for the *in vitro* quantitative detection of antibodies, including IgG, against the SARS-CoV-2 S protein RBD in human serum and plasma (26).

The objective of this multicenter European study was to evaluate the specificity and sensitivity of the Elecsys Anti-SARS-CoV-2 S immunoassay using pre-pandemic samples (from routine diagnostics or blood donation) and PCR-positive samples, respectively, as well as compare the performance of this quantitative test with other commercially available immunoassays in terms of specificity and sensitivity.

## Materials and methods

### Study design

The study was executed from August 17, 2020 to September 1, 2020 with samples tested at four European sites: Labor Augsburg MVZ GmbH, Augsburg, Germany; MVZ Labor Dr. Limbach & Kollegen GbR, Heidelberg, Germany; Interregionale Blutspende SRK AG (SRK Bern), Bern, Switzerland; and Krankenhaus Barmherzige Brüder, Regensburg, Germany. Samples were collected from those four sites, as well as from three additional sites: Labor Berlin – Charité Vivantes GmbH, Berlin, Germany; Tirol Kliniken, Innsbruck, Austria; and Deutsches Rotes Kreuz Blutspendedienst West, Hagen, Germany.

Samples from Augsburg and Heidelberg included those referred to the respective study site by physicians. Heidelberg also tested samples from employees and hospitalized patients, including a subset from patients receiving dialysis. All samples provided by the study site in Berlin were collected from hospitalized patients, including a subset from patients monitored in the intensive care unit (ICU). Samples tested in Regensburg were taken from employees and pediatric patients referred to the site by physicians.

These samples were collected and tested in accordance with applicable regulations, including relevant European Union directives and regulations, and the principles of the Declaration of Helsinki. All samples from Augsburg, Heidelberg, Berlin, and Hagen were anonymized. A statement was obtained from the Ethics Committee (EC) of the Landesärztekammer Bayern confirming that there are no objections to the use of anonymized leftover samples. From the internal EC at the study site in Bern (Switzerland) a waiver was received and from the internal EC at the study site in Innsbruck (Austria) an approval was received. For Regensburg (Germany), EC approvals were already in place, amendments were submitted to notify the EC about Elecsys Anti-SARS-CoV-2 S testing. At Augsburg, Heidelberg, and Bern the assays were performed on the cobas e 801 analyzer (Roche Diagnostics International Ltd, Rotkreuz, Switzerland), whereas at Regensburg the assays were performed on the cobas e 601 analyzer (Roche Diagnostics International Ltd, Rotkreuz, Switzerland).

### Serum and plasma samples

Anonymized frozen, residual samples from blood donation centers or routine laboratory diagnostics, as well as banked samples, were used for this study. For specificity analysis of the Elecsys Anti-SARS-CoV-2 S assay, 7880 presumed negative samples (5056 blood donor and 2824 diagnostic routine samples) that were collected before October 2019 were tested. The diagnostic routine cohort included samples from pregnancy screening and pediatrics. For the sensitivity analysis of the Elecsys Anti-SARS-CoV-2 S assay, 827 PCR-confirmed single or sequential samples from 272 different patients, with known time difference between blood draw and positive PCR test, were tested. Of these presumed negative and PCR-confirmed samples, 7903 were tested on the commercially available Elecsys^®^ Anti-SARS-CoV-2 assay (Roche Diagnostics International Ltd, Rotkreuz, Switzerland) (27). Additionally, a number of these samples were tested on other commercially available assays: LIAISON^®^ SARS-CoV-2 S1/S2 IgG (DiaSorin) (28), 2052 samples; EUROIMMUN Anti-SARS-CoV-2 IgG (29) and IgA assays (EUROIMMUN) (30), 1618 and 1624 samples, respectively; ARCHITECT SARS-CoV-2 IgG (Abbott) (31), 3068 samples; ADVIA Centaur^®^ SARS-CoV-2 Total (Siemens Healthineers) (32), 1064 samples; iFlash-SARS-CoV-2 IgG and IgM assays (Shenzhen YHLO Biotech Co) (33), both 1062 samples.

### Elecsys Anti-SARS-CoV-2 S assay

The Elecsys Anti-SARS-CoV-2 S immunoassay is a quantitative ECLIA that detects high-affinity antibodies to the SARS-CoV-2 S protein RBD and has a low risk of detecting weakly cross-reactive and unspecific antibodies. Results are automatically reported as the analyte concentration of each sample in U/mL, with <0.80 U/mL interpreted as negative for anti-SARS-CoV-2 S antibodies and ≥0.80 U/mL interpreted as positive for anti-SARS-CoV-2 S antibodies (Roche Diagnostics GmbH. Elecsys Anti-SARS-CoV-2 S assay method sheet. 2020; version 01) (26).

### Comparator assays

Specimens were analyzed using eight comparator immunoassays according to the manufacturer’s instructions. Interpretation of results was performed according to the manufacturer’s instructions.

The Elecsys Anti-SARS-CoV-2 assay is an ECLIA for the *in vitro* qualitative detection of antibodies, including IgG, against SARS-CoV-2, using a recombinant protein representing the nucleocapsid (N) antigen (27). Results are automatically calculated in the form of a cutoff index (COI), with COI values <1.0 interpreted as non-reactive (negative) for anti-SARS-CoV-2 N antibodies and ≥1.0 as reactive (positive) for anti-SARS-CoV-2 N antibodies (27).

The LIAISON SARS-CoV-2 S1/S2 IgG assay is an indirect chemiluminescence immunoassay (CLIA) for the quantitative detection of IgG anti-S1 and IgG anti-S2 antibodies to SARS-CoV-2 (28). Results are automatically calculated, with antibody concentrations expressed as arbitrary units (AU/mL). Concentrations of <12.0 AU/mL are interpreted as negative, ≥12.0 to <15.0 AU/mL are interpreted as equivocal, and ≥15.0 AU/mL are interpreted as positive (28). Equivocal values are referred to as ‘gray zone’ results.

The EUROIMMUN Anti-SARS-CoV-2 IgG and IgA assays are separate enzyme-linked immunosorbent assay (ELISAs) that detect IgG or IgA anti-S1 antibodies to SARS-CoV-2 (29, 30). Results are evaluated semi-quantitatively by calculation of a ratio in which the absorbance values of the controls or patient samples are related to the absorbance value of the calibrator (29, 30). For both assays, ratio results <0.8 are interpreted as negative, ≥0.8 to <1.1 are borderline, and ≥1.1 are positive (29, 30). Borderline values are referred to as ‘gray zone’ results.

The ARCHITECT SARS-CoV-2 IgG assay is a chemiluminescent microparticle immunoassay (CMIA) used for the qualitative detection of IgG antibodies against the N antigen (31). Results are expressed in signal-to-cutoff (S/CO) values, with <1.4 results interpreted as negative and ≥1.4 results interpreted as positive (31).

The ADVIA Centaur SARS-CoV-2 Total assay is a CLIA intended for the qualitative detection of antibodies against the RBD of the S1 protein (32). Results are reported in index values, with <1.0 interpreted as non-reactive (negative) for anti-SARS-CoV-2 antibodies and ≥1.0 interpreted as reactive (positive) for anti-SARS-CoV-2 antibodies (32).

The iFlash-SARS-CoV-2 IgM and IgG assays (33) are separate CLIAs used for the qualitative detection of IgM or IgG against the S and N proteins. The iFlash system automatically calculates the analytic concentration of each sample, with <10 AU/mL interpreted as non-reactive and ≥10 AU/mL interpreted as reactive for anti-SARS-CoV-2 IgM or IgG antibodies.

### Statistical analysis

Sample size estimations for specificity and sensitivity analyses were based on formulae proposed previously (34). Assuming specificities between 0.998 and 0.999 and a sensitivity of 0.999, samples sizes of 1698 to 20964 and 32 to 50 respectively, would be required to obtain a significance level of 0.05 and a power of 0.8. For specificity and sensitivity calculations, point estimates and two-sided 95% confidence intervals (CIs) using the exact method were computed employing R version 3.4.0 (35). In the sensitivity evaluation, assay results were assigned to the respective week after positive PCR result. In the comparison with other commercially available assays, only samples with paired measurements were included in the respective analyses. For the differences in estimated specificities and sensitivities between Elecsys Anti-SARS-CoV-2 S assay and the comparator assays, two-sided 95% Wald CIs were calculated as previously recommended (36). If these CIs did not include zero, differences were considered as statistically significant.

### Data availability

Qualified researchers may request access to individual patient level data through the clinical study data request platform (https://vivli.org/). Further details on Roche’s criteria for eligible studies are available here: https://vivli.org/members/ourmembers/. For further details on Roche’s Global Policy on the Sharing of Clinical Information and how to request access to related clinical study documents, see here: https://www.roche.com/research_and_development/who_we_are_how_we_work/clinical_trials/our_commitment_to_data_sharing.htm.

## Results

### Overall performance of the Elecsys Anti-SARS-CoV-2 S assay

#### Specificity in different target cohorts

Specificity of the Elecsys Anti-SARS-CoV-2 S assay was evaluated at three European sites (with samples from five European sites) using 7880 evaluable residual samples from blood donors and routine diagnostic testing; all of which were collected before October 2019 and presumed negative for SARS-CoV-2 antibodies. The overall specificity for all samples was 99.95% (95% CI: 99.87–99.99) (Table 1). There were four samples with reactive results of 1.790 U/mL, 0.900 U/mL, 0.870 U/mL, and 1.130 U/mL. Three of these reactive samples were from blood donor samples, of which one was collected in March 2016 (influenza season) at Innsbruck, Austria and two were collected in July/August 2018 (outside influenza season) at Bern, Switzerland (Table 1). There was no statistically significant difference in specificity between blood donor samples collected during or outside influenza season. The other reactive sample was from the pregnancy screening cohort in Augsburg (Table 1).

**Table 1.**
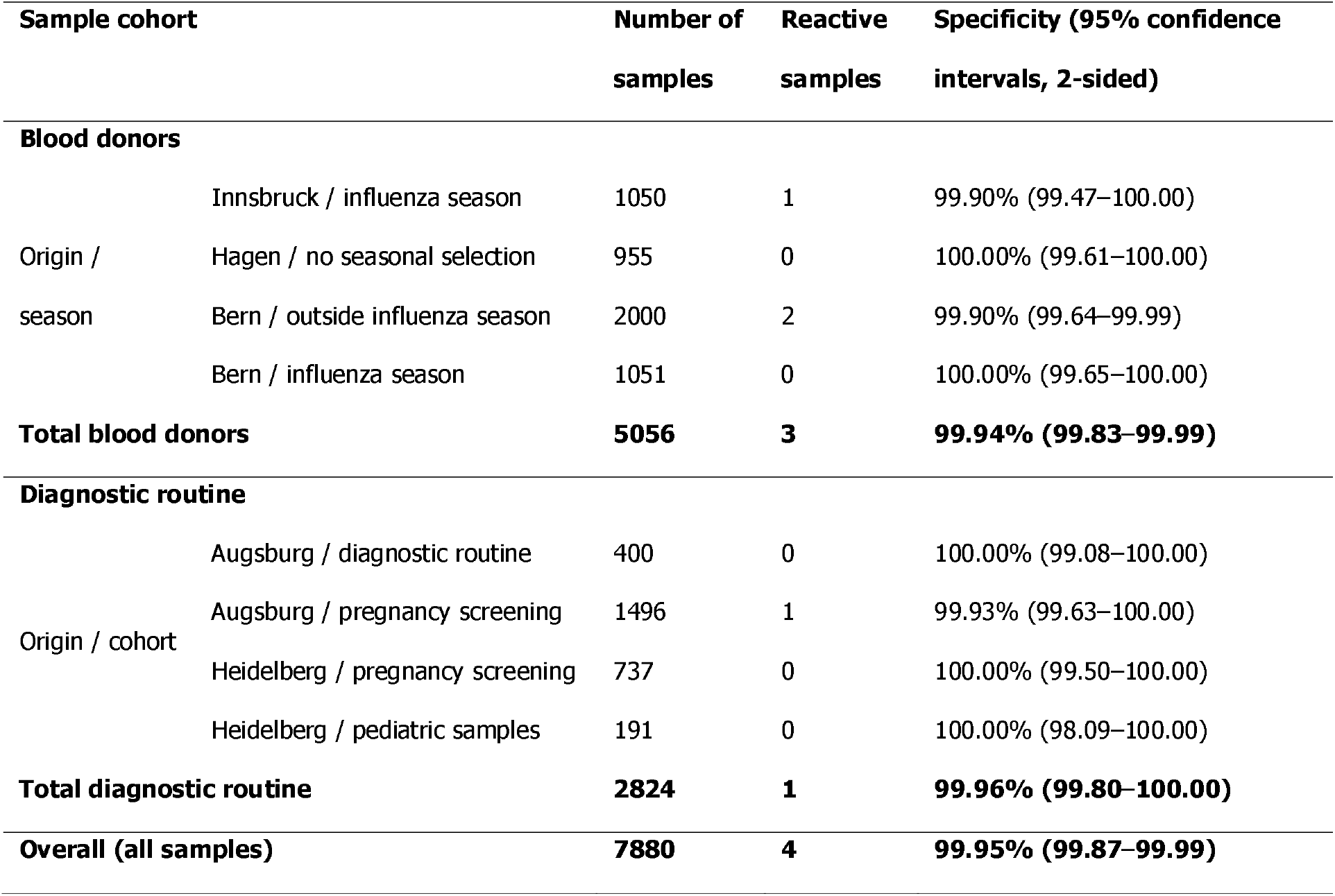
Specificity results for the Elecsys Anti-SARS-CoV-2 S assay.

| Sample cohort |  | Number of<br>samples | Reactive<br>samples | Specificity (95% confidence<br>intervals, 2-sided) |
| --- | --- | --- | --- | --- |
| **Blood donors** |  |  |  |  |
| Origin /<br>season | Innsbruck / influenza season | 1050 | 1 | 99.90% (99.47–100.00) |
|  | Hagen / no seasonal selection | 955 | 0 | 100.00% (99.61–100.00) |
|  | Bern / outside influenza season | 2000 | 2 | 99.90% (99.64–99.99) |
|  | Bern / influenza season | 1051 | 0 | 100.00% (99.65–100.00) |
| **Total blood donors** |  | **5056** | **3** | **99.94% (99.83–99.99)** |
| **Diagnostic routine** |  |  |  |  |
| Origin / cohort | Augsburg / diagnostic routine | 400 | 0 | 100.00% (99.08–100.00) |
|  | Augsburg / pregnancy screening | 1496 | 1 | 99.93% (99.63–100.00) |
|  | Heidelberg / pregnancy screening | 737 | 0 | 100.00% (99.50–100.00) |
|  | Heidelberg / pediatric samples | 191 | 0 | 100.00% (98.09–100.00) |
| **Total diagnostic routine** |  | **2824** | **1** | **99.96% (99.80–100.00)** |
| **Overall (all samples)** |  | **7880** | **4** | **99.95% (99.87–99.99)** |

#### Sensitivity in different target cohorts

In total, 827 evaluable single and sequential samples from 272 SARS-CoV-2 PCR-confirmed patients were evaluated at three European sites (with samples from four European sites). The time span of samples collected after positive PCR was between day 0 and day 120. For subjects with sequential blood draws with more than one sample per time interval, only the result of the last blood draw per given time interval was used for the respective sensitivity calculation. The sensitivity of the Elecsys Anti-SARS-CoV-2 S assay ≥14 days post-PCR (n=240) was 97.92% (95% CI: 95.21–99.32) (Table 2a). The resulting site-specific sensitivities for Augsburg, Berlin, Heidelberg, and Regensburg samples collected ≥14 days post-PCR confirmation were 100.00% (95% CI: 95.89–100.00), 100.00% (95% CI: 91.40– 100.00), 98.72% (95% CI: 93.06–99.97%), and 87.88% (95% CI: 71.80–96.60), respectively (Table 2b).

**Table 2a.** Overall sensitivity results for the Elecsys Anti-SARS-CoV-2 S assay.

| Days post-PCR-positive test | Number of samples | Reactive samples | Non-reactive samples | Sensitivity (95% confidence intervals, 2-sided) |
| --- | --- | --- | --- | --- |
| 0 to 6 | 44 | 25 | 19 | 56.82% (41.03–71.65) |
| 7 to 13 | 49 | 42 | 7 | 85.71% (72.76–94.06) |
| 14 to 20 | 47 | 46 | 1 | 97.87% (88.71–99.95) |
| 21 to 27 | 58 | 58 | 0 | 100.00% (93.84–100.00) |
| 28 to 34 | 43 | 43 | 0 | 100.00% (91.78–100.00) |
| 35 to 41 | 57 | 56 | 1 | 98.25% (90.61–99.96) |
| 42 to 48 | 47 | 47 | 0 | 100.00% (92.45–100.00) |
| 49 to 55 | 39 | 35 | 4 | 89.74% (75.78–97.13) |
| 56 to 62 | 33 | 33 | 0 | 100.00% (89.42–100.00) |
| ≥63 (up to 120) | 21 | 21 | 0 | 100.00% (83.89–100.00) |
| **All samples ≥14 (up to 120)<sup>1</sup>** | **240** | **235** | **5** | **97.92% (95.21–99.32)** |

<sup>1</sup> Only the last sample taken ≥14 days post-PCR of each patient is included in the sensitivity calculation.
PCR, polymerase chain reaction

**Table 2b.**
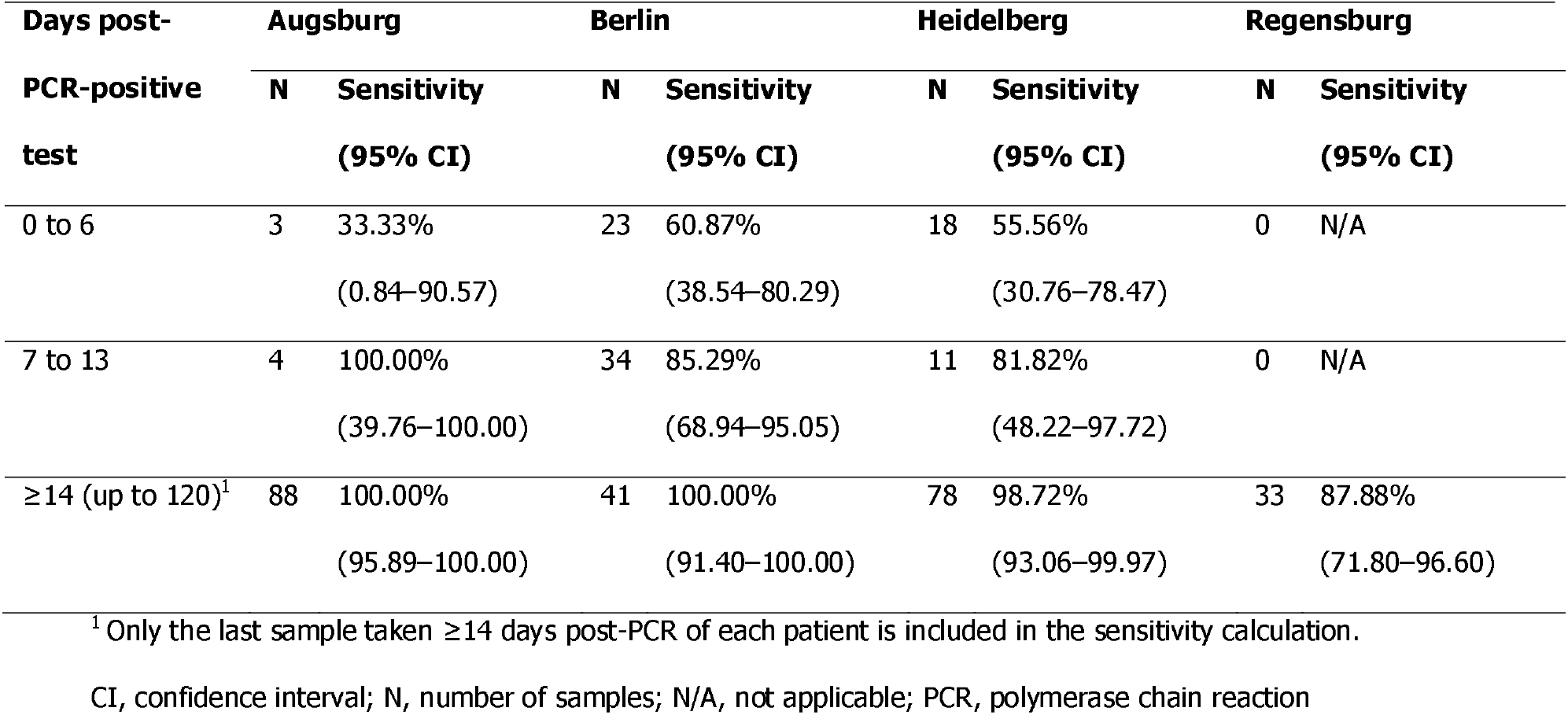
Sensitivity results for the Elecsys Anti-SARS-CoV-2 S assay per site.

#### Visualization of seroconversions and/or titer visualization

For all subjects with at least two sequential blood draws, trajectories were plotted to visualize antibody titer development from day 0 to 78 post-PCR-positive test (Figure 1). Most trajectories show a rapid increase in antibody titer and no considerable decline of antibody titer can be seen for the early and later blood draws. Once detected reactive, none of the subsequent samples drawn per subject showed a decline of titer below the cutoff.

**Figure 1.**
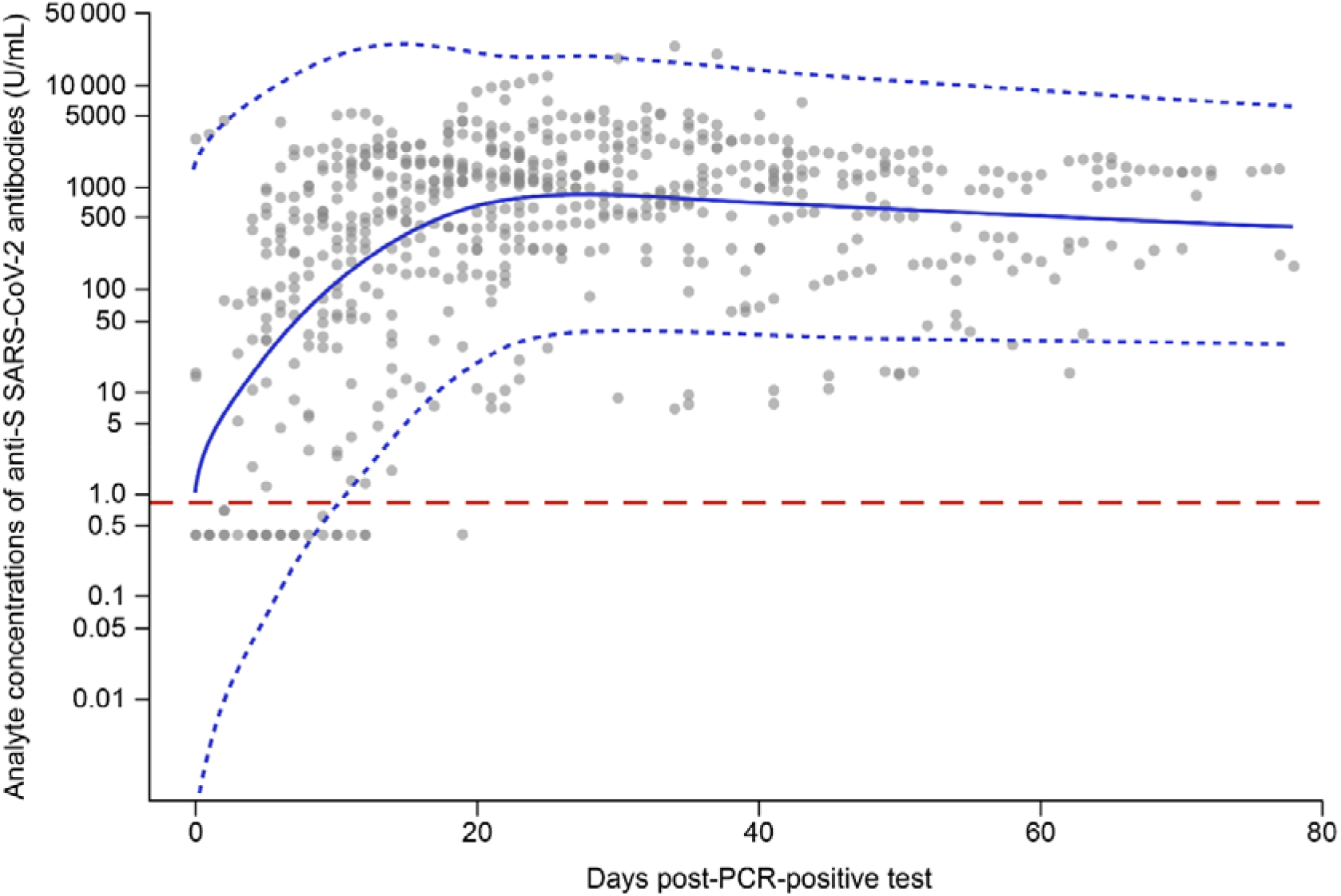
Longitudinal antibody titers of all subjects. Concentration of anti-S SARS-CoV-2 antibodies, as measured by the Elecsys Anti-S SARS-CoV-2 S immunoassay, over time (days 0 to 78) in sequential samples from all study sites. Each grey circle represents a different data point, with darker circles representing overlapping data points. The solid blue line represents the combined curve and the blue dashed lines represent the upper and lower confidence limits. The red dashed line indicates the assay cutoff limit (0.80 U/mL).

### Comparison with Elecsys Anti-SARS-CoV-2 assay

A direct method comparison between the Elecsys Anti-SARS-CoV-2 S assay and the commercially available Elecsys Anti-SARS-CoV-2 assay was performed. This included a total of 7903 samples comprising both confirmed positive samples from sensitivity testing and presumed positive samples with at least one positive antibody result (n=1011), as well as presumed negative samples from specificity testing cohort samples (n= 6892: n=4068 blood donors; n=2824 routine diagnostic). For all samples, the overall percent agreement (OPA) between the Elecsys Anti-SARS-CoV-2 S assay and the Elecsys Anti-SARS-CoV-2 assay was 99.30% (95% CI: 99.10–99.48) (Table S1a). For presumed negative samples and confirmed positive samples, the negative percent agreement (NPA) and positive percent agreement (PPA), respectively, between the S- and N-assays were >99% (Table S1b–c).

### Comparison with other commercially available assays

The performance of the Elecsys Anti-SARS-CoV-2 S immunoassay was compared with seven other commercially available SARS-CoV-2 assays, and sensitivity and specificity results, along with percent agreement, were recorded.

The OPA between the Elecsys Anti-SARS-CoV-2 S assay and other comparator tests was recorded (Table S2). The Elecsys Anti-SARS-CoV-2 S test had the highest OPA with the ARCHITECT SARS-CoV-2 IgG (N-assay), at 99.19% (95% CI: 98.80–99.47), and the lowest OPA with the ADVIA Centaur SARS-CoV-2 Total (S-assay), at 88.25% (95% CI: 86.16– 90.13) (Table S2).

### Specificity

The specificity of the Elecsys Anti-SARS-CoV-2 S assay was comparable or higher than the specificity of all tested comparator assays (Table 3). The specificity of the Elecsys Anti-SARS-CoV-2 S test was significantly higher compared with the LIAISON SARS-CoV-2 S1/S2 IgG, ADVIA Centaur SARS-CoV-2 Total, ARCHITECT SARS-CoV-2 IgG, iFlash-SARS-CoV-2 IgM, and EUROIMMUN Anti-SARS-CoV-2 IgG and IgA assays (Table S3a). No statistically significant difference was observed between the specificity of the Elecsys Anti-SARS-CoV-2 S assay compared with the iFlash-SARS-CoV-2 IgG assay (Table S3a).

**Table 3.** Specificity of the Elecsys Anti-SARS-CoV-2 S assay and other comparator assays.

| Assay | Number of samples | Reactive samples | Non-reactive samples | Specificity (95% confidence intervals, 2-sided) |
| --- | --- | --- | --- | --- |
| **LIAISON SARS-CoV-2 S1/S2 IgG** (GZ excl.)<sup>1</sup> | 2033 | 24 | 2009 | 98.82% (98.25–99.24) |
| **Elecsys Anti-SARS-CoV-2 S** |  | 1 | 2032 | 99.95% (99.73–100.00) |
| **LIAISON SARS-CoV-2 S1/S2 IgG** (GZ+) | 2040 | 31 | 2009 | 98.48% (97.85–98.97) |
| **Elecsys Anti-SARS-CoV-2 S** |  | 1 | 2039 | 99.95% (99.73–100.00) |
| **ADVIA Centaur SARS-CoV-2 Total** | 928 | 121 | 807 | 86.96% (84.62–89.06) |
| **Elecsys Anti-SARS-CoV-2 S** |  | 0 | 928 | 100.00% (99.60–100.00) |
| **ARCHITECT SARS-CoV-2 IgG** | 2932 | 9 | 2923 | 99.69% (99.42–99.86) |
| **Elecsys Anti-SARS-CoV-2 S** |  | 1 | 2931 | 99.97% (99.81–100.00) |
| **EUROIMMUN Anti-SARS-CoV-2 IgG** (GZ excl.) | 903 | 23 | 880 | 97.45% (96.20–98.38) |
| **Elecsys Anti-SARS-CoV-2 S** |  | 0 | 903 | 100.00% (99.59–100.00) |
| **EUROIMMUN Anti-SARS-CoV-2 IgG** (GZ+) | 924 | 44 | 880 | 95.24% (93.66–96.52) |
| **Elecsys Anti-SARS-CoV-2 S** |  | 0 | 924 | 100.00% (99.60–100.00) |
| **EUROIMMUN Anti-SARS-CoV-2 IgA** (GZ excl.) | 895 | 38 | 857 | 95.75% (94.22–96.98) |
| **Elecsys Anti-SARS-CoV-2 S** |  | 0 | 895 | 100.00% (99.59–100.00) |
| **EUROIMMUN Anti-SARS-CoV-2 IgA** (GZ+) | 928 | 71 | 857 | 92.35% (90.45–93.98) |
| **Elecsys Anti-SARS-CoV-2 S** |  | 0 | 928 | 100.00% (99.60–100.00) |
| **iFlash-SARS-CoV-2 IgG** | 928 | 0 | 928 | 100.00% (99.60–100.00) |
| **Elecsys Anti-SARS-CoV-2 S** |  | 0 | 928 | 100.00% (99.60–100.00) |
| **iFlash-SARS-CoV-2 IgM** | 928 | 4 | 924 | 99.57% (98.90–99.88) |
| **Elecsys Anti-SARS-CoV-2 S** |  | 0 | 928 | 100.00% (99.60–100.00) |

<sup>1</sup>For antibody assays with a gray zone, two calculations were performed. In the first calculation, all gray zone results were excluded from the analysis (GZ excl.) and in the second calculation these results were interpreted as reactive (GZ+).
GZ, gray zone

### Sensitivity

The sensitivity of the Elecsys Anti-SARS-CoV-2 S assay for detecting seropositive results was compared with six comparator assays; analysis compared with the LIAISON SARS-CoV-2 S1/S2 IgG test could not be performed due to a small sample size. Sensitivity was recorded for samples collected between 0–6, 7–13, and ≥14 days post-PCR-positive test. The sensitivity of the Elecsys Anti-SARS-CoV-2 S assay was equal to or higher than the sensitivity for all tested IgM, IgG, and total antibody assays at all time intervals (Table 4). The EUROIMMUN Anti-SARS-CoV-2 IgA assay showed a higher sensitivity in the 0–6 and 7–13 days post-PCR time intervals and a lower sensitivity in the ≥14 days post-PCR time interval compared with the Elecsys Anti-SARS-CoV-2 S assay (Table 4). The sensitivity of the Elecsys Anti-SARS-CoV-2 S assay at detecting antibodies ≥14 days post-PCR was significantly higher compared with the ARCHITECT SARS-CoV-2 IgG, iFlash-SARS-CoV-2 IgG and IgM, and EUROIMMUN Anti-SARS-CoV-2 IgG assays (Table S3b).

**Table 4.**
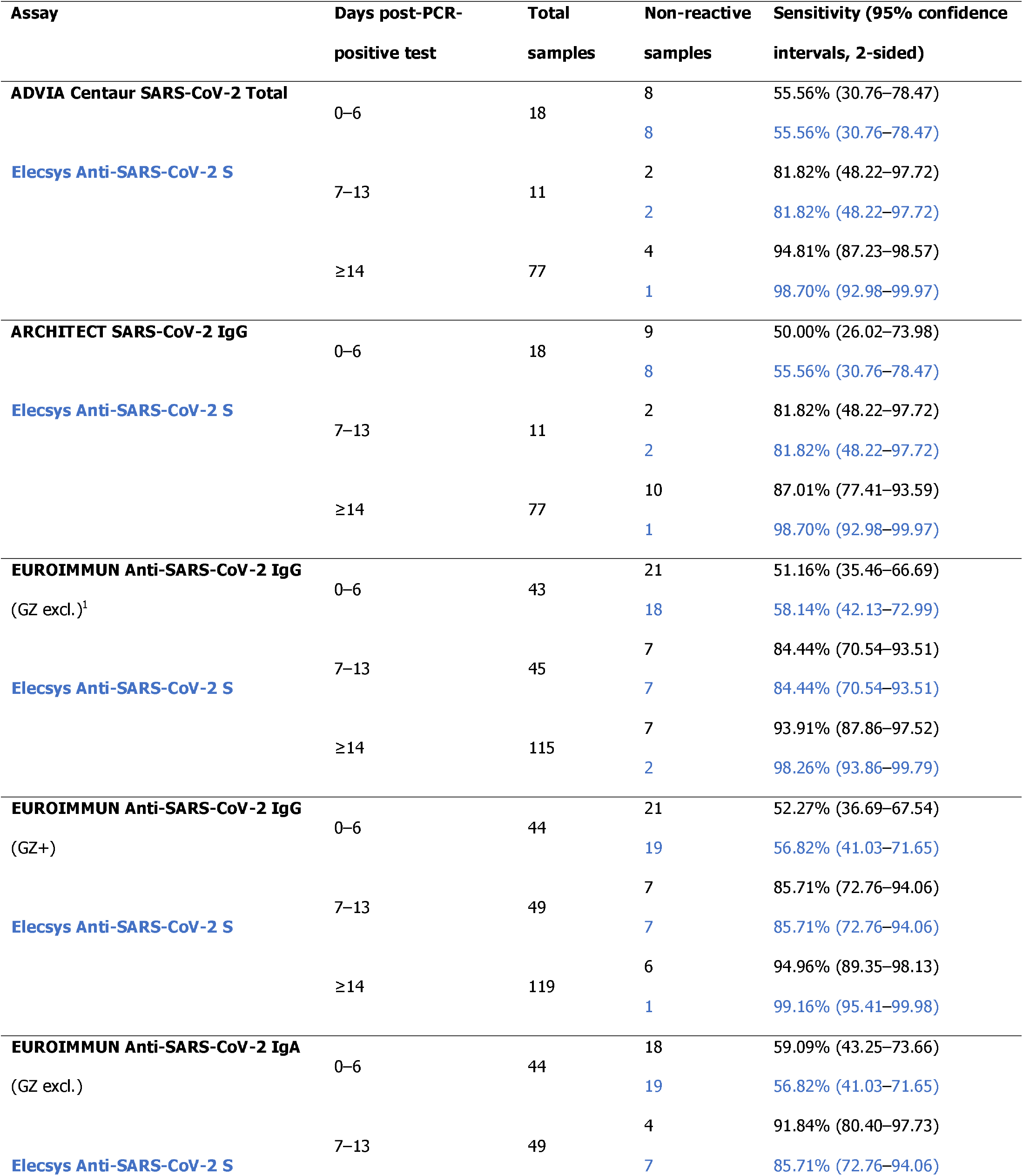
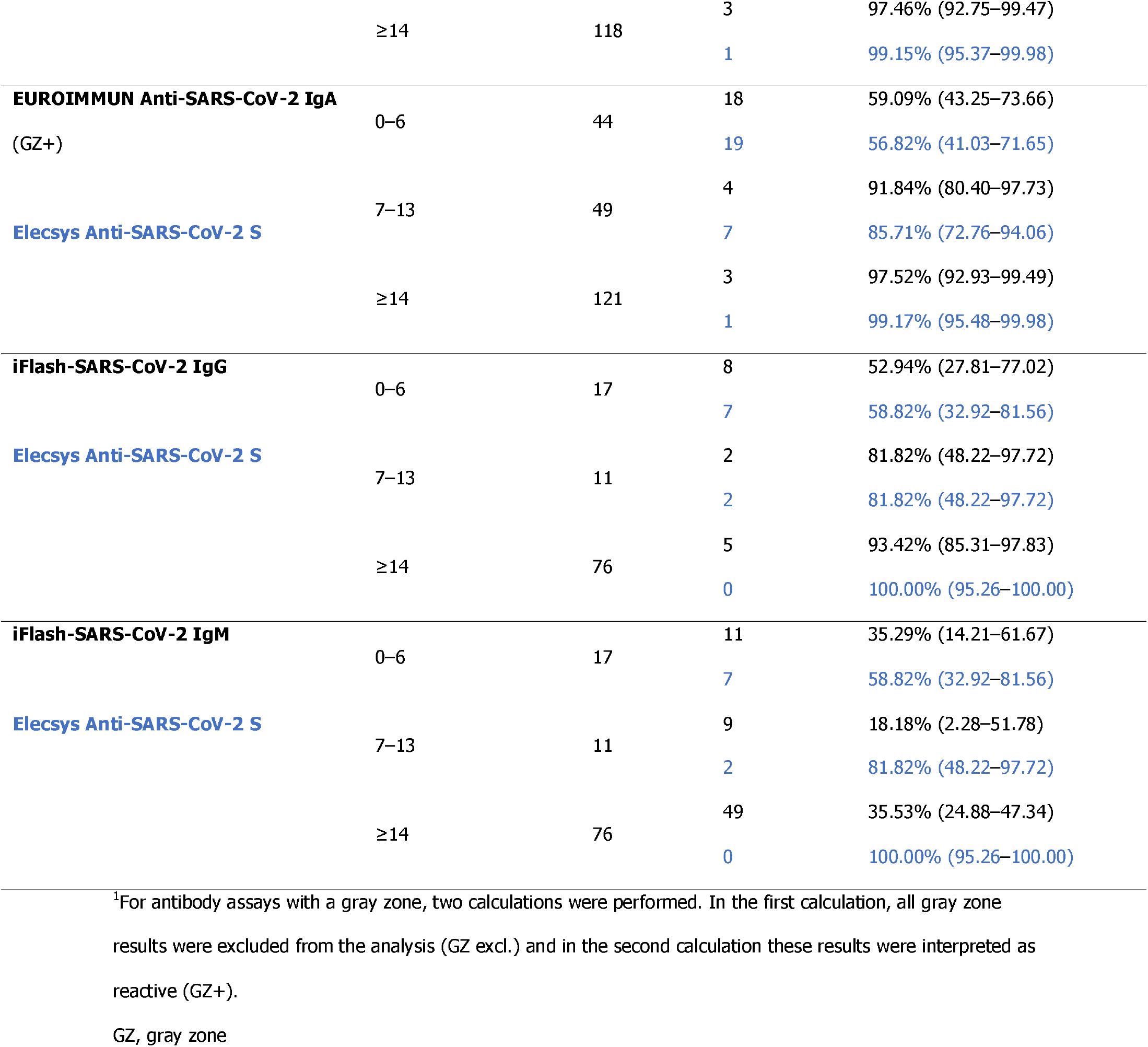
Sensitivity of the Elecsys Anti-SARS-CoV-2 S assay and other comparator assays.

## Discussion

Due to the COVID-19 pandemic, there is a pressing need to develop highly specific and sensitive serology tests to assist with the diagnosis of, and to reveal past exposure to, the SARS-CoV-2 virus (17), as well as to support the development of vaccines through distinguishing natural infection-induced immunity from vaccine-induced immunity (9, 18). This was the first multicenter study to demonstrate the performance of the automated Elecsys Anti-SARS-CoV-2 S immunoassay, which detects antibodies against the SARS-CoV-2 S protein RBD. Antibodies against the RBD have previously been shown to correlate strongly with protective neutralizing antibodies (37).

The results from our study revealed that the Elecsys Anti-SARS-CoV-2 S immunoassay displays a robust and favorable performance under routine conditions at multiple sites in Europe, with a very high specificity (99.95%) and sensitivity (97.92%) for the detection of anti-S antibodies. The point estimates for specificity and sensitivity are comparable to the values reported in the package insert of the Elecsys Anti-SARS-CoV-2 S assay (99.98% and 98.8%, respectively) (26). In addition, the Elecsys Anti-SARS-CoV-2 S assay showed a performance comparable with the commercially available Elecsys Anti-SARS-CoV-2 (N-assay), with 95% CIs that overlap (99.69–99.88% for specificity and 97.0–100% for sensitivity); both assays had a very high overall percent agreement. The Elecsys Anti-SARS-CoV-2 assay has a previously reported specificity and sensitivity ≥14 days post-confirmation of 99.8% and 99.5%, respectively (38).

The overall specificity of >99.9% determined in this study demonstrated that the Elecsys Anti-SARS-CoV-2 S is a highly specific assay for the detection of antibodies against SARS-CoV-2. Notably, this analysis included 2424 samples from pregnant women and pediatric populations. The availability of an accurate SARS-CoV-2 serology assay is particularly important for the pregnant population, considering the changes in the immune system that occur during pregnancy, which may increase the woman’s susceptibility to severe infection (39). Additionally, an antibody assay with a high specificity is imperative to reduce the risk of false-positive results, which may inaccurately indicate a past SARS-CoV-2 infection (40). The Elecsys Anti-SARS-CoV-2 S immunoassay demonstrated good specificity and sensitivity in direct comparison with other commercially available assays; both performance measurements were equal to or greater than those for all other evaluated comparator assays. These other assays have also been assessed in previous studies (17, 41-44). However, it is important to note that, for a direct comparison of sensitivity, the available assays differ with respect to assay designs (e.g. antibody classes used) as well as the targets (anti-N and anti-S) that they detect.

A multicenter comparison of seven serology assays, including the Elecsys Anti-SARS-CoV-2 assay, revealed a subpopulation of PCR-confirmed SARS-CoV-2 individuals who were persistently seronegative, which represents a proportion of patients that may be at risk for re-infection (45). Within the group of PCR-confirmed samples in our study, for which there was at least one S and one N antibody result from another comparator assay, there were samples from five patients all of which had a reactive Elecsys Anti-SARS-CoV-2 S test result, a reactive or ‘gray zone’ EUROIMMUN test result and a reactive ADVIA Centaur test result (S-assays), and a non-reactive Elecsys Anti-SARS-CoV-2 and ARCHITECT SARS-CoV-2 IgG test result (N-assays). There was only one sample that was commonly non-reactive in all S-based assays and reactive in all N-based assays. However, the blood draw was taken very early, at the same day as the PCR was done, and the follow-up sample was reactive with the Elecsys Anti-SARS-CoV-2 S assay. Data indicating differences in the kinetics of serology assays are ambiguous and there is little difference in the timing of these responses (42, 46-49). However, differences between N- and S- antigen-based assays should be taken into consideration when interpreting results.

A major strength of this study is the large cohort of presumed negative samples from multiple sites used to determine the specificity of the assay, as well as the multiple method comparison analyses performed and the various population cohorts used, with samples from pediatric and pregnant woman. Further studies should be performed to determine the sensitivity of the Elecsys Anti-SARS-CoV-2 S immunoassay on a larger sample group.

## Conclusion

This study demonstrated the performance of the Elecsys Anti-SARS-CoV-2 S immunoassay, with a very high specificity of 99.95% and sensitivity of 97.92% in samples ≥14 days post-PCR confirmation, which was equal to or greater than the performance of seven other commercially available immunoassays. Therefore, these data support the use of the Elecsys Anti-SARS-CoV-2 S immunoassay for reliable identification of past exposure to SARS-CoV-2 in various populations, and highlight the potential for the use of this assay in determining immune status during vaccine efficacy studies.

## Acknowledgements

The authors would like to acknowledge: Kathrin Schoenfeld (Roche Diagnostics) for her role in study conceptualization, study management, interpretation of analysis, and further critical input; Michael Laimighofer (Roche Diagnostics) for his role in database generation and data validation, statistical analysis plan, and formal analysis; and Sigrid Reichhuber and Janina Edion (Roche Diagnostics) for their role in investigational site management, data acquisition, and study monitoring. Medical writing support was provided by Katie Farrant, Elements Communications Ltd, Westerham, UK and was funded by Roche Diagnostics International Ltd, Rotkreuz, Switzerland. ELECSYS and COBAS E are trademarks of Roche Diagnostics. All other product names and trademarks are the property of their respective owners.

## References

1. Chan JF, Yuan S, Kok KH, To KK, Chu H, Yang J, Xing F, Liu J, Yip CC, Poon RW, Tsoi HW, Lo SK, Chan KH, Poon VK, Chan WM, Ip JD, Cai JP, Cheng VC, Chen H, Hui CK, Yuen KY. 2020. A familial cluster of pneumonia associated with the 2019 novel coronavirus indicating person-to-person transmission: a study of a family cluster. Lancet 395:514–523.

2. Wu F, Zhao S, Yu B, Chen YM, Wang W, Song ZG, Hu Y, Tao ZW, Tian JH, Pei YY, Yuan ML, Zhang YL, Dai FH, Liu Y, Wang QM, Zheng JJ, Xu L, Holmes EC, Zhang YZ. 2020. A new coronavirus associated with human respiratory disease in China. Nature 579:265–269.

3. World Health Organization. 2020. WHO Director-General’s remarks at the media briefing on 2019-nCoV on 11 February 2020. https://www.who.int/dg/speeches/detail/who-director-general-s-remarks-at-the-media-briefing-on-2019-ncov-on-11-february-2020. Accessed February 26 2021.

4. Naqvi AAT, Fatima K, Mohammad T, Fatima U, Singh IK, Singh A, Atif SM, Hariprasad G, Hasan GM, Hassan MI. 2020. Insights into SARS-CoV-2 genome, structure, evolution, pathogenesis and therapies: Structural genomics approach. Biochim Biophys Acta Mol Basis Dis 1866:165878.

5. Walls AC, Park YJ, Tortorici MA, Wall A, McGuire AT, Veesler D. 2020. Structure, function, and antigenicity of the SARS-CoV-2 spike glycoprotein. Cell 181:281–292 e6.

6. Tang T, Bidon M, Jaimes JA, Whittaker GR, Daniel S. 2020. Coronavirus membrane fusion mechanism offers a potential target for antiviral development. Antiviral Res 178:104792.

7. Wrapp D, Wang N, Corbett KS, Goldsmith JA, Hsieh CL, Abiona O, Graham BS, McLellan JS. 2020. Cryo-EM structure of the 2019-nCoV spike in the prefusion conformation. Science 367:1260–1263.

8. Ou X, Liu Y, Lei X, Li P, Mi D, Ren L, Guo L, Guo R, Chen T, Hu J, Xiang Z, Mu Z, Chen X, Chen J, Hu K, Jin Q, Wang J, Qian Z. 2020. Characterization of spike glycoprotein of SARS-CoV-2 on virus entry and its immune cross-reactivity with SARS-CoV. Nat Commun 11:1620.

9. Galipeau Y, Greig M, Liu G, Driedger M, Langlois MA. 2020. Humoral responses and serological assays in SARS-CoV-2 infections. Front Immunol 11:610688.

10. Liu W, Liu L, Kou G, Zheng Y, Ding Y, Ni W, Wang Q, Tan L, Wu W, Tang S, Xiong Z, Zheng S. 2020. Evaluation of nucleocapsid and spike protein-based enzyme-linked immunosorbent assays for detecting antibodies against SARS-CoV-2. J Clin Microbiol 58.

11. Guo L, Ren L, Yang S, Xiao M, Chang, Yang F, Dela Cruz CS, Wang Y, Wu C, Xiao Y, Zhang L, Han L, Dang S, Xu Y, Yang QW, Xu SY, Zhu HD, Xu YC, Jin Q, Sharma L, Wang L, Wang J. 2020. Profiling early humoral response to diagnose novel coronavirus disease (COVID-19). Clin Infect Dis 71:778–785.

12. To KK, Tsang OT, Leung WS, Tam AR, Wu TC, Lung DC, Yip CC, Cai JP, Chan JM, Chik TS, Lau DP, Choi CY, Chen LL, Chan WM, Chan KH, Ip JD, Ng AC, Poon RW, Luo CT, Cheng VC, Chan JF, Hung IF, Chen Z, Chen H, Yuen KY. 2020. Temporal profiles of viral load in posterior oropharyngeal saliva samples and serum antibody responses during infection by SARS-CoV-2: an observational cohort study. Lancet Infect Dis 20:565–574.

13. Long QX, Liu BZ, Deng HJ, Wu GC, Deng K, Chen YK, Liao P, Qiu JF, Lin Y, Cai XF, Wang DQ, Hu Y, Ren JH, Tang N, Xu YY, Yu LH, Mo Z, Gong F, Zhang XL, Tian WG, Hu L, Zhang XX, Xiang JL, D.HX, Liu HW, Lang CH, Luo XH, Wu SB, Cui XP, Zhou Z, Zhu MM, Wang J, Xue CJ, Li XF, Wang L, Li ZJ, Wang K, Niu CC, Yang QJ, Tang XJ, Zhang Y, Liu XM, Li JJ, Zhang DC, Zhang F, Liu P, Yuan J, Li Q, Hu JL, Chen J, et al. 2020. Antibody responses to SARS-CoV-2 in patients with COVID-19. Nat Med 26:845–848.

14. Zhao J, Yuan Q, Wang H, Liu W, Liao X, Su Y, Wang X, Yuan J, Li T, Li J, Qian S, Hong C, Wang F, Liu Y, Wang Z, He Q, Li Z, He B, Zhang T, Fu Y, Ge S, Liu L, Zhang J, Xia N, Zhang Z. 2020. Antibody responses to SARS-CoV-2 in patients with novel coronavirus disease 2019. Clin Infect Dis 71:2027–2034.

15. Lou B, Li TD, Zheng SF, Su YY, Li ZY, Liu W, Yu F, Ge SX, Zou QD, Yuan Q, Lin S, Hong CM, Yao XY, Zhang XJ, Wu DH, Zhou GL, Hou WH, Li TT, Zhang YL, Zhang SY, Fan J, Zhang J, Xia NS, Chen Y. 2020. Serology characteristics of SARS-CoV-2 infection after exposure and post-symptom onset. Eur Respir J 56.

16. Young BE, Ong SWX, Ng LFP, Anderson DE, Chia WN, Chia PY, Ang LW, Mak TM, Kalimuddin S, Chai LYA, Pada S, Tan SY, Sun L, Parthasarathy P, Fong SW, Chan YH, Tan CW, Lee B, Rötzschke O, Ding Y, Tambyah P, Low JGH, Cui L, Barkham T, Lin RTP, Leo YS, Renia L, Wang LF, Lye DC. 2020. Viral dynamics and immune correlates of COVID-19 disease severity. Clin Infect Dis doi:10.1093/cid/ciaa1280.

17. Kontou PI, Braliou GG, Dimou NL, Nikolopoulos G, Bagos PG. 2020. Antibody tests in detecting SARS-CoV-2 infection: A meta-analysis. Diagnostics (Basel) 10:319.

18. Ernst E, Wolfe P, Stahura C, Edwards KA. 2021. Technical considerations to development of serological tests for SARS-CoV-2. Talanta 224:121883.

19. Zhu FC, Guan XH, Li YH, Huang JY, Jiang T, Hou LH, Li JX, Yang BF, Wang L, Wang WJ, Wu SP, Wang Z, Wu XH, Xu JJ, Zhang Z, Jia SY, Wang BS, Hu Y, Liu JJ, Zhang J, Qian XA, Li Q, Pan HX, Jiang HD, Deng P, Gou JB, Wang XW, Wang XH, Chen W. 2020. Immunogenicity and safety of a recombinant adenovirus type-5-vectored COVID-19 vaccine in healthy adults aged 18 years or older: a randomised, double-blind, placebo-controlled, phase 2 trial. Lancet 396:479–488.

20. Widge AT, Rouphael NG, Jackson LA, Anderson EJ, Roberts PC, Makhene M, Chappell JD, Denison MR, Stevens LJ, Pruijssers AJ, McDermott AB, Flach B, Lin BC, Doria-Rose NA, O’Dell S, Schmidt SD, Neuzil KM, Bennett H, Leav B, Makowski M, Albert J, Cross K, Edara VV, Floyd K, Suthar MS, Buchanan W, Luke CJ, Ledgerwood JE, Mascola JR, Graham BS, Beigel JH. 2021. Durability of Responses after SARS-CoV-2 mRNA-1273 Vaccination. N Engl J Med 384:80–82.

21. World Health Organization. 2021. The COVID-19 candidate vaccine landscape. https://www.who.int/publications/m/item/draft-landscape-of-covid-19-candidate-vaccines. Accessed February 09 2021.

22. The New York Times. 2021. Coronavirus vaccine tracker. https://www.nytimes.com/interactive/2020/science/coronavirus-vaccine-tracker.html. Accessed February 09 2021.

23. Dai L, Gao GF. 2020. Viral targets for vaccines against COVID-19. Nat Rev Immunol doi:10.1038/s41577-020-00480-0.

24. Forni G, Mantovani A. 2021. COVID-19 vaccines: where we stand and challenges ahead. Cell Death Differ doi:10.1038/s41418-020-00720-9:1-14.

25. Ni L, Ye F, Cheng ML, Feng Y, Deng YQ, Zhao H, Wei P, Ge J, Gou M, Li X, Sun L, Cao T, Wang P, Zhou C, Zhang R, Liang P, Guo H, Wang X, Qin CF, Chen F, Dong C. 2020. Detection of SARS-CoV-2-specific humoral and cellular immunity in COVID-19 convalescent individuals. Immunity 52:971–977 e3.

26. Roche Diagnostics GmbH. 2020. Elecsys® Anti-SARS-CoV-2 S assay method sheet; V1. https://diagnostics.roche.com/global/en/products/params/elecsys-anti-sars-cov-2-s.html. Accessed February 26 2021.

27. Roche Diagnostics GmbH. 2020. Elecsys® Anti-SARS-CoV-2 assay method sheet; 09203095501 V6.0. https://www.fda.gov/media/137605/download. Accessed February 26 2021.

28. DiaSorin. 2020. LIAISON® SARS-CoV-2 S1/S2 IgG assay method sheet; M0870004366/D 09/20. https://www.diasorin.com/sites/default/files/allegati_prodotti/covid_-_brochure_igg_unica_m0870004366-d_low.pdf. Accessed February 26 2021. 29.

29. EUROIMMUN. 2020. Anti-SARS-CoV-2 ELISA (IgG) product data sheet; EI_2606_D_UK_A06, 10/2020. https://www.coronavirus-diagnostics.com/documents/Indications/Infections/Coronavirus/EI_2606_D_UK_A.pdf. Accessed February 26 2021.

30. EUROIMMUN. 2020. Anti-SARS-CoV-2 ELISA (IgA) product data sheet; EI_2606_D_UK_B04, 10/2020. https://www.coronavirus-diagnostics.com/documents/Indications/Infections/Coronavirus/EI_2606_D_UK_B.pdf. Accessed February 26 2021.

31. Abbott. 2020. ARCHITECT SARS-CoV-2 IgG assay method sheet. https://www.fda.gov/media/137383/download. Accessed February 26 2021.

32. Siemens. 2020. ADVIA Centaur® SARS-CoV-2 Total (COV2T) assay method sheet; 11206904_EN Rev. 01, 2020-05. https://www.fda.gov/media/138446/download. Accessed February 26 2021.

33. Shenzhen YHLO Biotech Co Ltd. 2020. iFlash-SARS-CoV-2 IgM/IgG antibody test product overview. http://en.szyhlo.com/cmscontent/iFlash-SARS-CoV-2-IgMIgG-Antibody-Test-439.html. Accessed February 26 2021.

34. Hajian-Tilaki K. 2014. Sample size estimation in diagnostic test studies of biomedical informatics. J Biomed Inform 48:193–204.

35. R Core Team. 2017. R: A language and environment for statistical computing. R Foundation for Statistical Computing, Vienna, Austria. https://www.R-project.org/. Accessed February 26 2021.

36. Wenzel D, Zapf A. 2013. Difference of two dependent sensitivities and specificities: Comparison of various approaches. Biom J 55:705–718.

37. Premkumar L, Segovia-Chumbez B, Jadi R, Martinez DR, Raut R, Markmann A, Cornaby C, Bartelt L, Weiss S, Park Y, Edwards CE, Weimer E, Scherer EM, Rouphael N, Edupuganti S, Weiskopf D, Tse LV, Hou YJ, Margolis D, Sette A, Collins MH, Schmitz J, Baric RS, de Silva AM. 2020. The receptor binding domain of the viral spike protein is an immunodominant and highly specific target of antibodies in SARS-CoV-2 patients. Sci Immunol 5.

38. Muench P, Jochum S, Wenderoth V, Ofenloch-Haehnle B, Hombach M, Strobl M, Sadlowski H, Sachse C, Torriani G, Eckerle I, Riedel A. 2020. Development an validation of the Elecsys Anti-SARS-CoV-2 immunoassay as a highly specific tool for determining past exposure to SARS-CoV-2. J Clin Microbiol 58.

39. Dashraath P, Wong JLJ, Lim MXK, Lim LM, Li S, Biswas A, Choolani M, Mattar C, Su LL. 2020. Coronavirus disease 2019 (COVID-19) pandemic and pregnancy. Am J Obstet Gynecol 222:521–531.

40. Farnsworth CW, Anderson NW. 2020. SARS-CoV-2 serology: much hype, little data. Clin Chem 66:875–877.

41. Meyer B, Torriani G, Yerly S, Mazza L, Calame A, Arm-Vernez I, Zimmer G, Agoritsas T, Stirnemann J, Spechbach H, Guessous I, Stringhini S, Pugin J, Roux-Lombard P, Fontao L, Siegrist CA, Eckerle I, Vuilleumier N, Kaiser L, Geneva Center for Emerging Viral D. 2020. Validation of a commercially available SARS-CoV-2 serological immunoassay. Clin Microbiol Infect 26:1386–1394.

42. Kohmer N, Westhaus S, Ruhl C, Ciesek S, Rabenau HF. 2020. Brief clinical evaluation of six high-throughput SARS-CoV-2 IgG antibody assays. J Clin Virol 129:104480.

43. Okba NMA, Muller MA, Li W, Wang C, GeurtsvanKessel CH, Corman VM, Lamers MM, Sikkema RS, de Bruin E, Chandler FD, Yazdanpanah Y, Le Hingrat Q, Descamps D, Houhou-Fidouh N, Reusken C, Bosch BJ, Drosten C, Koopmans MPG, Haagmans BL. 2020. Severe acute respiratory syndrome coronavirus 2-specific antibody responses in coronavirus disease patients. Emerg Infect Dis 26:1478–1488.

44. Ocmant A, Roisin S, De Meuter R, Brauner J. 2021. Clinical performance of the Advia Centaur anti-SARS-CoV-2 chemiluminescent immunoassay related to antibody kinetics. J Med Virol doi:10.1002/jmv.26800.

45. Oved K, Olmer L, Shemer-Avni Y, Wolf T, Supino-Rosin L, Prajgrod G, Shenhar Y, Payorsky I, Cohen Y, Kohn Y, Indenbaum V, Lazar R, Geylis V, Oikawa MT, Shinar E, Stoyanov E, Keinan-Boker L, Bassal R, Reicher S, Yishai R, Bar-Chaim A, Doolman R, Reiter Y, Mendelson E, Livneh Z, Freedman LS, Lustig Y. 2020. Multi-center nationwide comparison of seven serology assays reveals a SARS-CoV-2 non-responding seronegative subpopulation. EClinicalMedicine 29:100651.

46. Burbelo PD, Riedo FX, Morishima C, Rawlings S, Smith D, Das S, Strich JR, Chertow DS, Davey RT, Jr., Cohen JI. 2020. Detection of nucleocapsid antibody to SARS-CoV-2 is more sensitive than antibody to spike protein in COVID-19 patients. medRxiv doi:10.1101/2020.04.20.20071423.

47. Wang Y, Zhang L, Sang L, Ye F, Ruan S, Zhong B, Song T, Alshukairi AN, Chen R, Zhang Z, Gan M, Zhu A, Huang Y, Luo L, Mok CKP, Al Gethamy MM, Tan H, Li Z, Huang X, Li F, Sun J, Zhang Y, Wen L, Li Y, Chen Z, Zhuang Z, Zhuo J, Chen C, Kuang L, Wang J, Lv H, Jiang Y, Li M, Lin Y, Deng Y, Tang L, Liang J, Huang J, Perlman S, Zhong N, Zhao J, Malik Peiris JS, Li Y, Zhao J. 2020. Kinetics of viral load and antibody response in relation to COVID-19 severity. J Clin Invest 130:5235–5244.

48. Van Elslande J, Decru B, Jonckheere S, Van Wijngaerden E, Houben E, Vandecandelaere P, Indevuyst C, Depypere M, Desmet S, Andre E, Van Ranst M, Lagrou K, Vermeersch P. 2020. Antibody response against SARS-CoV-2 spike protein and nucleoprotein evaluated by four automated immunoassays and three ELISAs. Clin Microbiol Infect 26:1557 e1–1557 e7.

49. Johnson M, Wagstaffe HR, Gilmour KC, Mai AL, Lewis J, Hunt A, Sirr J, Bengt C, Grandjean L, Goldblatt D. 2020. Evaluation of a novel multiplexed assay for determining IgG levels and functional activity to SARS-CoV-2. J Clin Virol 130:104572.

