## Supplemental Table 1, Table 2 and Table 3 for "Performance evaluation of the Roche Elecsys Anti-SARS-CoV-2 S immunoassay"

**Supplemental Material**

**Table S1a. Agreement of the Elecsys Anti-SARS-CoV-2 assay compared with the Elecsys Anti-SARS-CoV-2 S assay for presumed negative and confirmed positive samples**

|  | **Elecsys Anti-SARS-CoV-2** | | **Total** |
| --- | --- | --- | --- |
| **Elecsys Anti-SARS-CoV-2 S** | **Reactive samples** | **Non-reactive samples** |  |
| **Reactive** | 878 | 40 | 918 |
| **Non-reactive** | 15 | 6970 | 6985 |
| **Total** | 893 | 7010 | **7903** |
| **OPA (95% CI)** |  |  | 99.30% (99.10–99.48) |

CI, confidence interval; OPA, overall percent agreement

**Table S1b. Agreement of the Elecsys Anti-SARS-CoV-2 assay compared with the Elecsys Anti-SARS-CoV-2 S assay for presumed negative samples**

|  | **Elecsys Anti-SARS-CoV-2** | | **Total** |
| --- | --- | --- | --- |
| **Elecsys Anti-SARS-CoV-2 S** | **Reactive samples** | **Non-reactive samples** |  |
| **Reactive** | 0 | 4 | 4 |
| **Non-reactive** | 10 | 6878 | 6888 |
| **Total** | 10 | 6882 | **6892** |
| **NPA (95% CI)** |  | 99.94% (99.85–99.98) |  |
| **OPA (95% CI)** |  |  | 99.80% (99.66–99.89) |

CI, confidence interval; NPA, negative percent agreement; OPA, overall percent agreement

**Table S1c. Agreement of the Elecsys Anti-SARS-CoV-2 assay compared with the Elecsys Anti-SARS-CoV-2 S assay for confirmed positive samples**

|  | **Elecsys Anti-SARS-CoV-2** | | **Total** |
| --- | --- | --- | --- |
| **Elecsys Anti-SARS-CoV-2 S** | **Reactive samples** | **Non-reactive samples** |  |
| **Reactive** | 878 | 36 | 914 |
| **Non-reactive** | 5 | 92 | 97 |
| **Total** | 883 | 128 | **1011** |
| **PPA (95% CI)** | 99.43% (98.68–99.82) |  |  |
| **OPA (95% CI)** |  |  | 95.94% (94.54–97.07) |

CI, confidence interval; OPA, overall percent agreement; PPA, positive percent agreement

**Table S2. Overall percent agreement of the Elecsys Anti-SARS-CoV-2 S assay and other comparator assays**

| **Comparator assay** | **Cohort** | **Number of samples**^1^ | | **OPA (95% CI)** |
| --- | --- | --- | --- | --- |
| **LIAISON SARS-CoV-2 S1/S2 IgG** | Specificity | 2040 | 2052 | 98.20% (97.52–98.73) |
|  | Sensitivity | 12 |  |  |
| **ADVIA Centaur SARS-CoV-2 Total** | Specificity | 928 | 1064 | 88.25% (86.16–90.13) |
|  | Sensitivity | 136 |  |  |
| **ARCHITECT SARS-CoV-2 IgG** | Specificity | 2932 | 3068 | 99.19% (98.80–99.47) |
|  | Sensitivity | 136 |  |  |
| **EUROIMMUN Anti-SARS-CoV-2 IgG** | Specificity | 924 | 1618 | 95.18% (94.02–96.17) |
|  | Sensitivity | 694 |  |  |
| **EUROIMMUN Anti-SARS-CoV-2 IgA** | Specificity | 928 | 1624 | 94.27% (93.03–95.35) |
|  | Sensitivity | 696 |  |  |
| **iFlash-SARS-CoV-2 IgG** | Specificity | 928 | 1062 | 99.15% (98.40–99.61) |
|  | Sensitivity | 134 |  |  |
| **iFlash-SARS-CoV-2 IgM** | Specificity | 928 | 1062 | 92.00% (90.20–93.56) |
|  | Sensitivity | 134 |  |  |

^1^Number of samples tested over all sites with a corresponding Elecsys Anti-SARS-CoV-2 S result.

CI, confidence interval; OPA, overall percent agreement

**Table S3a. Evaluation of the differences in specificities between the Elecsys Anti-SARS-CoV-2 S assay and other comparator assays**

| **Comparator assay** | **Difference in specificity (95% Wald-CI)** | **Significant** |
| --- | --- | --- |
| **LIAISON SARS-CoV-2 S1/S2 IgG** (GZ excl.)^1^ | 1.13% (0.65–1.61) | Yes |
| **LIAISON SARS-CoV-2 S1/S2 IgG** (GZ+)^1^ | 1.47% (0.93–2.01) | Yes |
| **ADVIA Centaur SARS-CoV-2 Total** | 13.04% (10.87–15.21) | Yes |
| **ARCHITECT SARS-CoV-2 IgG** | 0.27% (0.06–0.48) | Yes |
| **EUROIMMUN Anti-SARS-CoV-2 IgG** (GZ excl.)^1^ | 2.55% (1.52–3.57) | Yes |
| **EUROIMMUN Anti-SARS-CoV-2 IgG** (GZ+)^1^ | 4.76% (3.39–6.14) | Yes |
| **EUROIMMUN Anti-SARS-CoV-2 IgA** (GZ excl.)^1^ | 4.25% (2.92–5.57) | Yes |
| **EUROIMMUN Anti-SARS-CoV-2 IgA** (GZ+)^1^ | 7.65% (5.94–9.36) | Yes |
| **iFlash-SARS-CoV-2 IgG** | 0.00% (0.00–0.00) | No |
| **iFlash-SARS-CoV-2 IgM** | 0.43% (0.01–0.85) | Yes |

^1^For antibody assays with a gray zone, two calculations were performed. In the first calculation, all gray zone results were excluded from the analysis (GZ excl.) and in the second calculation these results were interpreted as reactive (GZ+).

CI, confidence interval; GZ, gray zone

**Table S3b. Evaluation of the differences in sensitivities (only for time interval ≥14 days after PCR) between the Elecsys Anti-SARS-CoV-2 S assay and other comparator assays**

| **Comparator assay** | **Difference in sensitivity (95% Wald-CI)** | **Significant** |
| --- | --- | --- |
| **ADVIA Centaur SARS-CoV-2 Total** | 3.90% (-0.43–8.22) | No |
| **ARCHITECT SARS-CoV-2 IgG** | 11.69% (4.51–18.86) | Yes |
| **EUROIMMUN Anti-SARS-CoV-2 IgG** (GZ excl.)^1^ | 4.35% (0.62–8.08) | Yes |
| **EUROIMMUN Anti-SARS-CoV-2 IgG** (GZ+)^1^ | 4.20% (0.60–7.81) | Yes |
| **EUROIMMUN Anti-SARS-CoV-2 IgA** (GZ excl.)^1^ | 1.69% (-0.63–4.02) | No |
| **EUROIMMUN Anti-SARS-CoV-2 IgA** (GZ+)^1^ | 1.65% (-0.62–3.92) | No |
| **iFlash-SARS-CoV-2 IgG** | 6.58% (1.01–12.15) | Yes |
| **iFlash-SARS-CoV-2 IgM** | 64.47% (53.71–75.23) | Yes |

^1^For antibody assays with a gray zone, two calculations were performed. In the first calculation, all gray zone results were excluded from the analysis (GZ excl.) and in the second calculation these results were interpreted as reactive (GZ+).

CI, confidence interval; GZ, gray zone
